# Educational attainment reduces the risk of suicide attempt among individuals with and without psychiatric disorders independent of cognition: a multivariable Mendelian randomization study with more than 815,000 participants

**DOI:** 10.1101/2019.12.14.19014787

**Authors:** Daniel B. Rosoff, Zachary A. Kaminsky, Falk W. Lohoff

**Affiliations:** Section on Clinical Genomics and Experimental Therapeutics, National Institute on Alcohol Abuse and Alcoholism, National Institutes of Health, Bethesda, MD, USA; Royal’s Institute of Mental Health Research, University of Ottawa, Ottawa, Canada

**Keywords:** educational attainment, cognitive performance, suicide attempt, Mendelian randomization, UK Biobank

## Abstract

**Background:** Rates of suicidal ideation, attempts and completions are increasing and identifying causal risk factors continues to be a public health priority. Observational literature has shown that educational attainment (EA) and cognitive performance (CP) can influence suicide attempt risk; however, due to residual confounding and reverse causation, the causal nature of these relationships is unknown.

**Methods:** We perform a multivariable two-sample Mendelian randomization (MR) analysis to disentangle the effects of EA and CP on suicide attempt risk. We use summary statistics from recent genome-wide association studies (GWAS) of EA, CP, household income versus suicide attempt risk in individuals with and without mental disorders, with more than 815,000 combined study participants.

**Results:** We found evidence that both EA and CP significantly reduced the risk of suicide attempt when considered separately in single variable MR (SVMR) (Model 1 EA odds ratio (OR), 0.524, 95% CI, 0.412-0.666, *P* = 1.07⨯10^−7^; CP OR, 0.714, 95% CI, 0.577-0.885, *P* = 0.002). When simultaneously analyzing EA,CA, and adjusting for household income but not comorbid mental disorders (Model 1), we found evidence that the direct effect of EA, independent of CP, on suicide attempt risk was greater than the total effect estimated by SVMR, with EA, independent of CP, significantly reducing the risk of suicide attempt by almost 66% (95% CI, 43%-79%); however, the effect of CP was no longer significant independent of EA (Model 1 EA OR, 0.342, 95% CI, 0.206-0.568, *P* = 1.61×10^−4^; CP OR, 1.182, 95% CI, 0.842-1.659, *P* = 0.333). Further, when accounting for comorbid mental disorders (Model 2), these results did not significantly change: we found EA significantly reduced the risk of suicide attempt by 55% (35%-68%), a lower point estimate but still within the 95% confidence interval of Model 1; the effect of CP was still not significant (Model 2 EA OR, 0.450, 95% CI, 0.314-0.644, *P* < 1.00×10^−4^; CP OR, 1.143, 95% CI, 0.803-1.627, *P* = 0.475).

**Conclusions:** Our results show that even after accounting for comorbid mental disorders and adjusting for household income, EA, but not CP, is a causal risk factor in suicide attempt. These findings could have important implications for health policy and prevention programs aimed at reducing the increasing rates of suicide.

## INTRODUCTION

Suicide is a leading cause of death with approximately 800,000 deaths per year worldwide [1], and the age-adjusted suicide rate in the United States has increased 33% from 1999 to 2017 with more than 120 Americans dying by suicide each day [2]. With an estimated 2.1 million discharges for self-inflicted injuries from emergency departments and acute care hospitals were reported in the US in 2013 alone [3] and $50.8 billion in medical expenditures and lost productivity [4], suicidal behavior is a major public health and economic burden [5, 6]. Given the incalculable emotional and psychological costs for families, friends, and the community at-large [6], the true public health burden of suicidality is difficult to estimate; however, these statistics highlight the importance of identifying causal risk factors for developing effective suicide reduction and prevention strategies [7].

While more than 90% of people who attempt or commit suicide have a psychiatric disorder [8], the pathways to suicidal behavior are complex, and involve the dynamic interaction of psychological, genetic, social, and cultural, factors [9]. For e.g., adverse childhood experiences, like sexual abuse and maladaptive parenting may contribute to suicidal behaviors, either directly [10], or through psychiatric disorders [9]. Further, the precise genetic basis of suicidal behavior remains largely unknown [9]; however, twin studies have suggested an approximate heritability of suicidal behavior of up to 55% [11], while adoption, twin and family studies have also shown that the genetic basis of suicidal behavior is, in part, distinct from that of psychiatric disorders [8, 12], and recent genome-wide association studies (GWAS) have identified the first genome-wide significant loci associated with suicide attempts in the general population [12] and psychiatric cohort [8].

Observational literature has shown that low educational attainment (EA) may be a major risk factor for suicidal behavior [13-16], which may contribute to the disparity in mortality across socioeconomic classes [17]. Further, this disparity due in part to suicide, may be growing with one study reporting that among adults 45-54, compared to college graduates, those with a high school degree or less, were 2.4 times more likely to commit suicide in 2013 while only 1.7 times more likely in 1999 [18]. However, caution is needed inferring causality from multivariable adjusted regression in observational data [19-21], and observational studies are subject to reverse causation and residual confounding [20, 22] from correlated factors like cognitive performance and other socioeconomic status variables, like income, that have also been shown to affect suicidal behavior [23, 24]. While randomized control trials (RCTs) are the “gold standard” of causal inference [25], it is impossible to create an RCT examining the effect of EA on suicidal behavior. However, robust evidence for causal relationship between EA and suicidal behavior is vital for the formulation and evaluation for the development of targeted prevention programs [26].

Mendelian randomization (MR) is a genetic epidemiology method that uses randomly inherited genetic variants as instrumental variables to assess possible causal relationships between environmental exposures (e.g., EA), and outcomes (e.g. suicidal behavior). As genetic variants are not modifiable by confounders and specific in their association with the exposure, MR is analogous to RCTs, where if the genetically-predicted values of the environmental exposure are associated with the outcome, then causal inference may be drawn from their association [20, 27], and is an important analytical strategy when RCTs are impractical or unethical [20]. Recent MR studies have found evidence for causal effects of EA on multiple behaviors and health outcomes, including alcohol consumption patterns and alcohol dependence [28], smoking [29, 30], physical activity [31], and cardiovascular disease [32-34].

While these studies suggest increasing years of education would address health disparities, the genetics of EA is complex [35], and failing to account for cognitive performance, which is heritable [36], strongly associated with EA [37, 38], and could be driving the observed protective effect of increased EA. Put another way, evaluating the independent effects of EA and cognitive performance is important for determining the relationships between education and disparities in health outcomes, including suicide. Multivariable MR (MVMR) is a method that allows for evaluation of separate, but correlated exposures like EA and cognitive performance [30, 39] by simultaneously estimating the effects of each exposure using a genetic instrument with potentially overlapping genetic variants [31]. Resulting MVMR estimates represent the direct effect of each exposure and can be interpreted of as evidence for a causal effect of one exposure given a constant level of the other exposure (i.e., the effect of EA holding cognitive performance constant).

In this study, we leveraged the largest publicly available GWAS on EA (N ≤ 766,345) [36], cognitive performance (N ≤ 257,828) [36], and hospital-based records of either a primary or secondary diagnosis of a suicide attempt (N ≤ 50,264) [40] and performed single-variable and MVMR analyses using genetic variants associated with EA and cognitive performance to evaluate the distinct effects on risk of suicide attempt in individuals with and without comorbid mental disorders. Given the strong genetic correlation of EA and cognitive performance with markers of socioeconomic status (SES) [41, 42], we included genetic variants associated with income (N ≤ 397,751) [43] in our final MVMR models.

## METHODS

### Study design and data sources

Figure 1 displays the overall design for the two-sample single-variable MR (SVMR) and MVMR analyses of the effect of EA and CP – adjusting for household income – on risk of suicide attempt of the study. We used publicly available summary statistics from three GWAS consortia (Table 1 in the Supplement; web links for downloading the data provided). All studies have existing ethical permissions from their respective institutional review boards and include participant informed consent and included rigorous quality control (Methods 1 in the Supplement). As all analyses herein are based on publicly available summary data, no ethical approval from an institutional review board was required.

**Table 1.** Single-Variable and Multivariable Inverse Variance Weighted Mendelian Randomization Associations Between Educational Attainment and Cognitive Performance on Risk of Suicide Attempt in Individuals With and Without Mental Disorders

| OUTCOMES: |  |  |  |  |  |  |  |  |  |  |
| --- | --- | --- | --- | --- | --- | --- | --- | --- | --- | --- |
| SUICIDE ATTEMPT: MODEL 1 |  |  |  |  |  | SUICIDE ATTEMPT: MODEL 2 |  |  |  |  |
| EXPOSURES | N SNPS | OR | OR LCI | OR UCI | p-value | N SNPS | OR | OR LCI | OR UCI | p-value |
| **SVMR:** |  |  |  |  |  |  |  |  |  |  |
| Educational Attainment | 223 | **0.524** | **0.412** | **0.666** | **1.07E-07** | 224 | **0.687** | **0.540** | **0.874** | **2.20E-03** |
| Cognitive Performance | 104 | **0.714** | **0.577** | **0.885** | **2.02E-03** | 104 | **0.794** | **0.636** | **0.990** | **4.01E-02** |
| Household Income | 41 | **0.591** | **0.389** | **0.901** | **1.41E-02** | 41 | 0.721 | 0.480 | 1.081 | 1.13E-01 |
| **MVMR:** |  |  |  |  |  |  |  |  |  |  |
| Educational Attainment | 288 | **0.450** | **0.314** | **0.644** | **1.00E-04** | 288 | **0.556** | **0.388** | **0.796** | **2.00E-03** |
| Cognitive Performance | 288 | 1.044 | 0.764 | 1.426 | 7.86E-01 | 288 | 1.081 | 0.789 | 1.482 | 6.29E-01 |
| **MVMR: Adjusting for Income** |  |  |  |  |  |  |  |  |  |  |
| Educational Attainment | 282 | **0.342** | **0.206** | **0.568** | **1.61E-04** | 283 | **0.437** | **0.258** | **0.739** | **2.00E-03** |
| Cognitive Performance | 282 | 1.182 | 0.842 | 1.659 | 3.33E-01 | 283 | 1.143 | 0.803 | 1.627 | 4.57E-01 |
| Household Income | 282 | 1.101 | 0.627 | 1.932 | 7.38E-01 | 283 | 1.225 | 0.683 | 2.197 | 4.96E-01 |

Results are presented as odds ratios (OR) with 95% confidence intervals for the effect of a unit increase in Educational Attainment (years schooling), standardized Cognitive Performance score, and categorical increase in average annual household income before tax, on the risk of suicide attempt (recorded non-fatal suicide attempt, including secondary diagnoses of poisoning by drugs or other substances, or injuries to hand, wrist, and forearm). Model 1 was based upon iPSYCH Suicide Attempt Risk GWAS not accounting for comorbid mental disorders (N=50,260); Model 2 was based upon iPSYCH Suicide Attempt Risk GWAS accounting for diagnosed comorbid mental disorders in same cohort sample (N=50,260): schizophrenia, bipolar disorder, affective disorders, autism spectrum disorder, anorexia, and “any other disorder”. Abbreviations: SVMR, single-variable Mendelian randomization; MVMR, multivariable Mendelian randomization; N, number; SNPs, single-nucleotide polymorphisms; OR, odds ratio; OR LCI, 95% confidence interval lower bound; OR UCI, 95% confidence interval upper bound.

**Figure 1.**
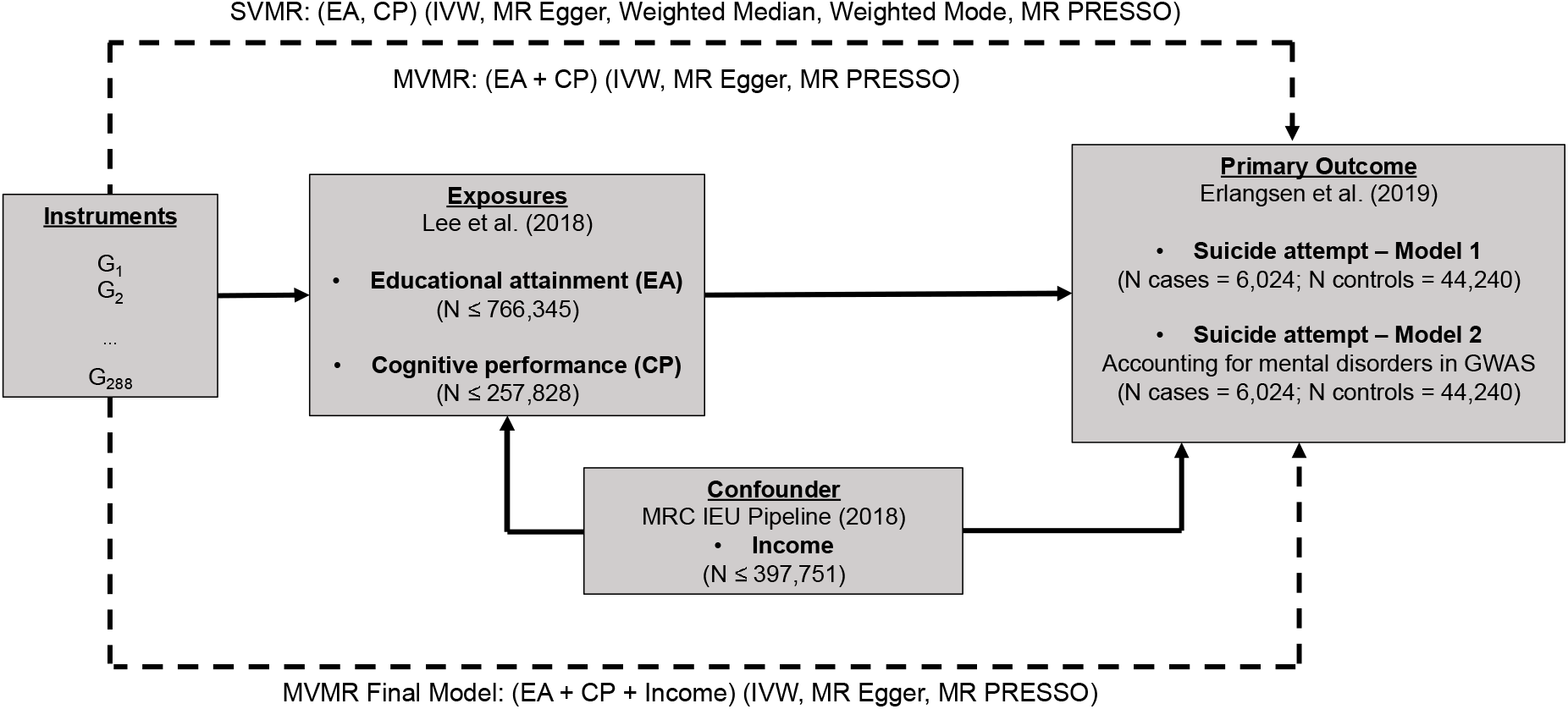
Study design schematic. Dashed lines indicate Mendelian randomization (MR) analyses of SVMR and MVMR exposure instruments on the outcome suicide attempt risk; MR methods/ sensitivity analyses in parentheses. Abbreviations: G, instrument variants; EA, educational attainment; CP, cognitive performance; Income, average household income before tax; MR, Mendelian randomization; MVMR, multivariable MR; SVMR, single-variable MR; IVW, inverse variance weighted MR; MR-PRESSO, Mendelian Randomization Pleiotropy Residual Sum and Outlier; SNP, single-nucleotide polymorphism.

### Instruments

We extracted SNPs for educational attainment (EA) from the recent SSGAC (Social Science Genetic Association Consortium) GWAS meta-analysis of 70 separate GWASs for a total of 766,345 individuals of European ancestry including 442,183 from the UK Biobank prospective cohort study collected across the United Kingdom from 2006-2010 [36]. Using the standard heuristic significance threshold for these dimensional MR studies to make the work manageable, we included all SNPs associated at genome-wide significance (P < 5×10^−8^) with the phenotype “educational attainment” (EA); EA was constructed from an imputed years-of education equivalent for each International Standard Classification of Education (ISCED) category mapped from each of cohort’s survey measures due to differences across countries in educational systems and qualifications. Across all cohorts (the 23andMe Cohort not included in the analyses herein), the sample-size weighted mean of EA is 16.8 years schooling (standard deviation 4.2 years). We pruned the results to exclude all SNPs with a pairwise linkage disequilibrium (LD) R^2^ > 0.001, giving us 318 independent SNPs associated with EA for the SVMR analyses (Table 2 in the Supplement).

We extracted SNPs for cognitive performance (CP) from the companion SSGAC GWAS meta-analysis of the Cognitive Genomics Consortium (COGENT) GWAS on general cognitive performance in 35,298 individuals combined with the GWAS of cognitive performance in 222,543 individuals of the UK Biobank prospective cohort study, both of predominantly European ancestry (total sample size 257,841). We included all SNPs associated with CP at genome-wide significance (P < 5 ⨯ 10^−8^). In the COGENT study, the phenotype was defined as the first principle component of three or more neuropsychological tests including but not limited to tests assessing digit span, digit symbol coding, phonemic fluency, semantic fluency, trail-making, verbal memory for stories, verbal memory for words, visual memory, vocabulary and word reading; in the UK Biobank, the standardized score on up to four tests taken by each respondent of a two-minute verbal-numerical assessment of fluid intelligence (comprised of thirteen logic and reasoning questions) was used for the phenotype: the genetic correlation between the score and general intelligence among children has been shown in prior studies to be approximately 0.8. We pruned the results to exclude all SNPs with a pairwise LD R^2^ > .001, leaving 157 independent SNPs associated with CP for SVMR analyses (Table 3 in the Supplement).

To account for income in our MVMR analyses, we extracted SNPs from the MRC-IEU UK Biobank GWAS based on the responses of 397,751 individuals of European ancestry including 442,183 from the UK Biobank prospective cohort study [43]. We included all SNPs associated at genome-wide significance (*P* < 5 ⨯ 10^−8^) with the PHESANT derived phenotype (UK Biobank data-field 738) from five categorically ordered responses of the study, with the first category “less than £18,000”, up the fifth category “more than £100,000”, categorizing average total household income before tax (Income). We pruned the results to exclude all SNPs with a pairwise linkage disequilibrium (LD) R^2^ > 0.001, leaving 48 independent SNPs associated with Income for the SVMR analyses (Table 4 in the Supplement).

For our MVMR analyses combining educational attainment and cognitive performance, we included all independent SNPs (LD R^2^ < .001, 10000 kb) which were genome-wide significant in either the GWASs of either EA or CP, deleting duplicate SNPs, resulting in an MVMR instrument comprised of 343 SNPs. Finally, adjusting for income, we combined those SNPs and the 41 independent SNPs for Income, deleting again duplicate SNPs, and pruning to exclude SNPs with a pairwise linkage LD R^2^ > .001, resulting in an MVMR (EA, CP and income, combined) of 340 SNPS.

### Suicide attempt outcomes

We extracted summary statistics on the associations of the instruments SNPs with suicide attempt (SA) from the recent iPSYCH GWASs in a sample constructed from the 1,472,762 singletons born in Denmark between 1981 and 2005, and residing in Denmark on their first birthday, including 57,377 diagnosed with one or more mental disorders, according to the 10^th^ Revision of International Classification of Diseases *i*.*e*. schizophrenia, bipolar disorders, affective disorders, autism spectrum disorder, anorexia, and, additionally, a non-psychiatric population-based random sample of 30,000 [35]. Sample participants were further screened by the Danish Psychiatric Central Research Register and National Registry of Patients for diagnoses of non-fatal suicide attempts; cases also included those with combinations of diagnoses with the main diagnoses recorded as a mental disorder, and a secondary diagnosis recorded as poisoning by drugs or other substances, or injuries to hand, wrist, and forearm, well-established proxies for suicide attempt [40]. The control group consisted of all persons not diagnosed with one or more suicide attempts with and without diagnosed mental disorders. After quality control, and excluding children, 50,264 remained in the sample for the GWAS, of which 6,024 (12%) recorded at least one suicide attempt, and 44,240 (88%) did not [35]. For the 6,024 cases, 17.9% were aged 15-19 years, 37.2% 20-24 years, 35.6% 25-29 years, and 9.4% 30-34 years; 54.7% of the cases had 1 recorded suicide attempt; 22.4%, 2; 14.8%, 3-4; 6.9%, 5-9; and 2.0%, 10+.

The primary iPSYCH GWAS (Model 1) did not account for comorbid mental disorders: the GWAS was conducted using logistic regression to calculate the log odds ratio of risk of suicide attempt with only gender, years followed, and first ten principal components of genetic ancestry included as covariates. The companion iPSYCH GWAS (Model 2) accounted for comorbid mental disorders by including a series of binary covariates for diagnoses of the following: schizophrenia, bipolar disorder, affective disorder, autism spectrum disorders, anorexia, and “any other disorder”. We adopt the Model 1 and 2 notation for the purposes of the present study.

### Extraction of instruments from outcomes GWAS and harmonization of effect alleles

Of the 318 possible genome-wide significant SNPs associated with EA, 270 SNPs were present in both iPSYCH Models 1 and 2 suicide attempt (SA) risk GWASs, and 46 SNPs were removed during harmonization for being palindromic with intermediate allele frequency, leaving 224 SNPs for SVMR analysis. Of the 157 possible SNPs associated with CA, 127 SNPs were present in the SA GWASs, and 23 SNPs were removed during harmonization for being palindromic with intermediate allele frequency, leaving 104 SNPs for SVMR analysis. Of the 343 possible independent SNPs of the combined MVMR instrument set for EA and CA, 288 SNPs were present in the SA GWASs, leaving 280 SNPs for MVMR analysis; and further, adjusting for income, 283 SNPs. We calculated the F statistics to assess the strength of the instruments (present in the SA GWAS), and all exceeded the threshold F-statistic = 10 recommended for MR analysis [44] (EA F-statistic = 59.802, 95% CI lower bound = 58.111; CP F-statistic = 55.234, 95% CI lower bound = 52.851; Income F-statistic = 45.265, 95% CI lower bound = 41.867).

### Sample overlap

Participant overlap in samples used to estimate genetic associations between exposure and outcome in two sample MR can bias results [39, 45], so we endeavor to minimize overlapping to reduce this source of weak instrument bias. In the instant study, we avoided overlap between cohorts used in the educational attainment, cognitive performance and income GWASs on the one hand, and the cohorts used in the iPSYCH suicide attempts cohorts on the other.

### Statistical analysis

For SVMR, applied to assess the total effects of EA and CP on risk of suicide attempt, with and without accounting for comorbid mental disorders, we used inverse-variance weighted MR (MR IVW) (single variable weighted linear regression) along with the complementary MR Egger method, to assess the evidence of the causal effects of educational attainment and cognitive performance on risk of suicide attempts, so as to detect the sensitivity of the results to different patterns of violations of IV assumptions: consistency of results across methods strengthens an inference of causality [46]. MR IVW is generally regarded as the main method: in the absence of pleiotropy and assuming the instruments are valid, MR IVW returns unbiased estimates of a causal effect are returned so long as horizontal pleiotropy is balanced [46, 47]. MR Egger extends MR IVW by not setting the intercept to zero, thus allowing the net-horizontal pleiotropic effect across all SNPs to be unbalanced or directional (i.e. some SNPs could be acting on the outcome through a pathway other than through the exposure) [47, 48]. MR Egger returns unbiased causal effect estimates even if the assumption of no horizontal pleiotropy is violated for all SNPs, but the estimates are less precise than MR IVW.

For MVMR, applied to assess the independent direct effects of EA and CP on risk for suicide attempts, with and without accounting for comorbid mental disorders, we used the extension developed by Burgess et al. (2015) of the inverse-variance weighted MR method, performing multivariable weighted linear regression (variants uncorrelated, random effect model) with the intercept term set to zero [47], and, additionally, the MVMR extension of the MR Egger method to correct for both measured and unmeasured pleiotropy [49].

### Sensitivity analyses and diagnostics

To evaluate heterogeneity in genetic instruments effects, indicating potential violations of the instrumental variable (IV) assumptions, we used the MR Egger intercept test [50], the Cochran Q heterogeneity test [51], and the MR <u>p</u>leiotropy <u>res</u>idual <u>s</u>um and <u>o</u>utlier (MR-PRESSO) test [52]. A MR Egger regression intercept is generally interpreted as the average pleiotropic effect across all instruments: MR-Egger regression thus provides a test for average pleiotropy, and further has been extended to correct for both measured and unmeasured pleiotropy in MVMR [49]. The Cochran Q test, used to identify outliers in regression analysis generally, has been applied in MR to detect average pleiotropy: pleiotropy can induce heterogeneity of individual ratio estimates [52]. MR-PRESSO detects pleiotropic bias in MR caused by violation of the exclusion restriction IV assumption; extending the principal of the Cochran Q test, MR-PRESSO provides a global test to detect pleiotropic bias and identify sources of the bias (so called outlier SNPs), and further has been extended to detect pleiotropic bias also in MVMR [52]. We used the MR-PRESSO global test to identify outlier SNPS; removing the outlier SNPs, we reran the MR, and retested so as to determine whether removing outliers resolved the detected heterogeneity.

## RESULTS

We generally looked for those estimates (1) substantially agreeing in direction and magnitude across complementary MR methods, (2) exceeding nominal significance in MR IVW, (3) not indicating bias from horizontal pleiotropy (MR-PRESSO global *P* > 0.01; and/or also MR Egger intercept *P* > 0.01), and, for SVMR, (4) indicating true causal effect directionality (Steiger directionality test *P* < 0.01). Complete MR results with test statistics are presented in Tables 5 and 6 in the Supplement. MR-PRESSO outlier corrected results are presented in Table 1 and Figures 2 and 3.

**Figure 2.**
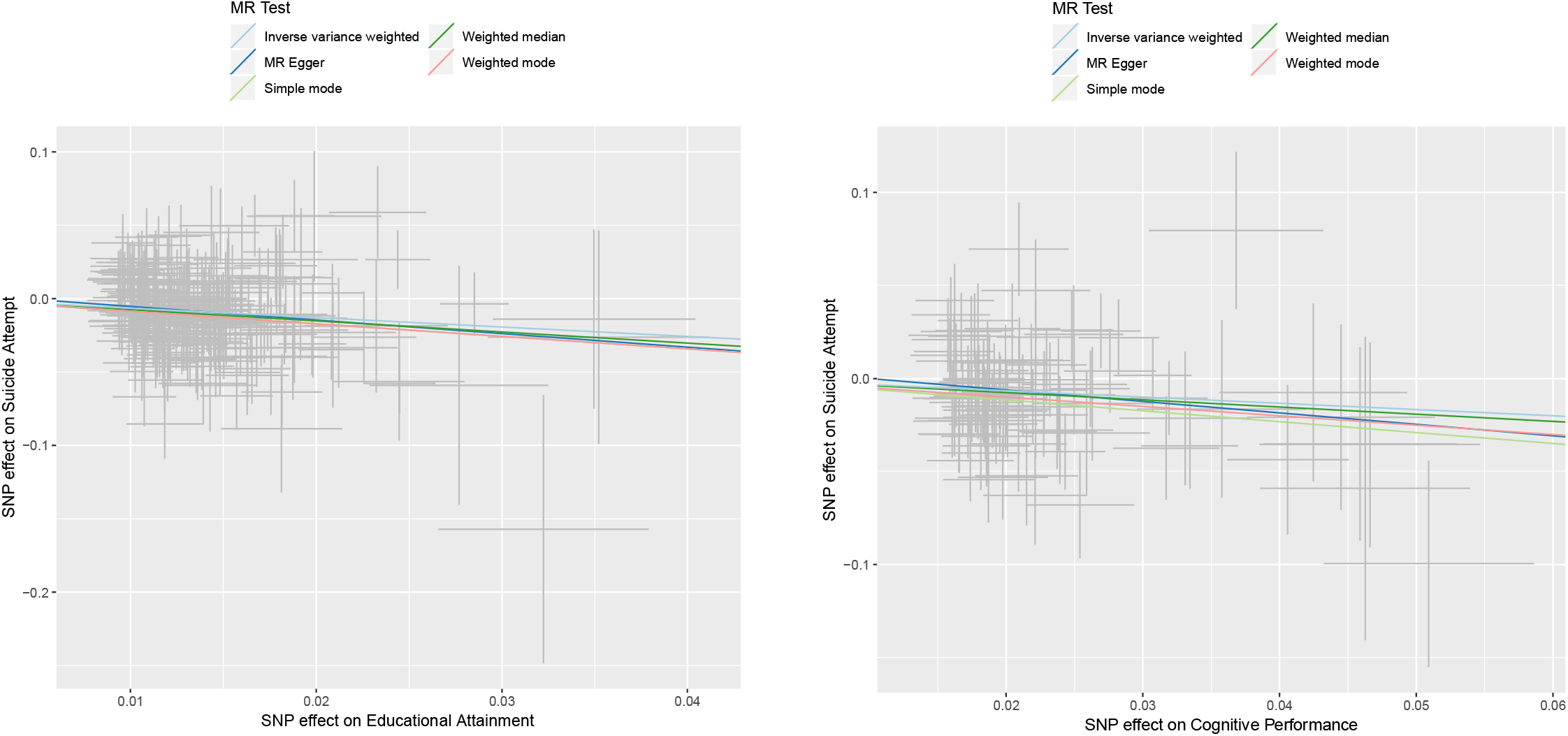
Single variable Mendelian randomization analysis for (LEFT) educational attainment and (RIGHT) cognitive performance on risk of suicide attempts. Abbreviations: MR, Mendelian randomization; and SNP, single-nucleotide polymorphism.

**Figure 3.**
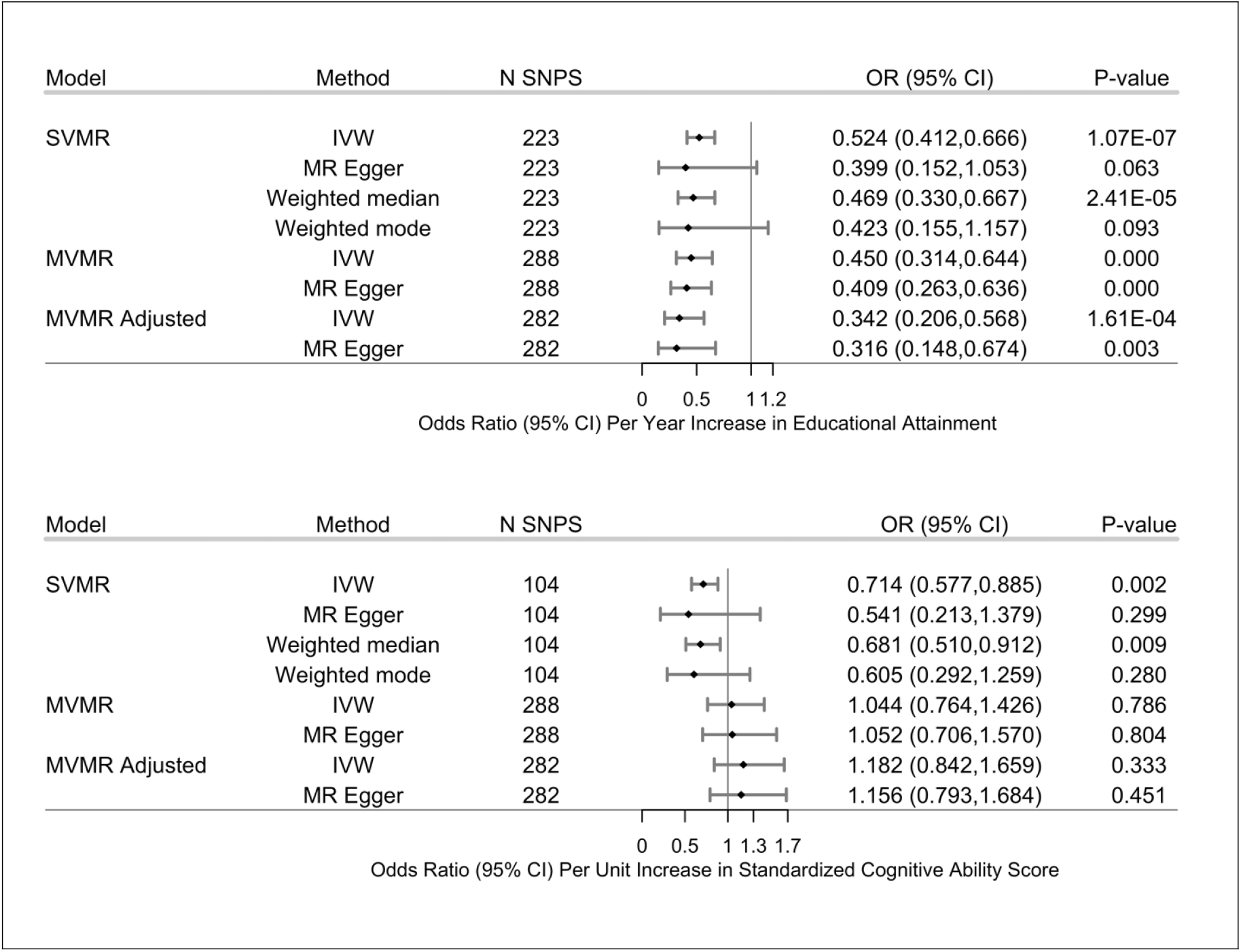
Effects of the genetic variants for increased educational attainment (TOP) and increased cognitive performance (BOTTOM) on risk of suicide attempt (Model 1). Instruments for each model were the genome wide significant (*P*□<□5□×□10^−8^) single nucleotide polymorphisms (SNPs) extracted from the educational attainment (EA) and cognitive performance (CP) GWASs, respectively for the SVMR analyses, combined for the MVMR analyses, that were independent at a linkage disequilibrium (LD) *R*^2^□=□0.001, with clumping distance□set at 10,000□kb, and found in the suicide attempt GWAS. Models are as follows: (1) SVMR results from inverse variance weighted (IVW) and three complementary two-sample MR methods, *i*.*e*. EA and CP analyzed separately: the estimates are considered to be the total effect (direct plus indirect effect) of the single exposure on the outcome; (2) MVMR results from IVW and MR Egger MV methods, with EA and CP analyzed simultaneously: the estimates are interpreted as the direct effect of the exposure on the outcome, independent of the effect of the other exposure; (3) MVMR adjusted: MVMR model adjusted for Average Household Income (Before Tax). All results shown are cleaned of variants identified as outliers by the MR PRESSO test (MR-PRESSO *P*□<□0.10). Cochran Q tests did not indicate heterogeneity and MR Egger intercept test did not indicate pleiotropy for any model. Error bars indicate 95% confidence intervals. EA, educational attainment; CP, cognitive performance; MR, Mendelian randomization; SVMR, single variable MR; MVMR, multivariable MR; IVW, inverse-variance weighted MR; OR, odds ratio; CI, confidence interval.

### SVMR

We used SVMR to assess the total effects of each exposure on our two models of risk of suicide attempt (Models 1 and 2; Model 2 using summary statistics from the SA GWAS accounting for comorbid mental disorders). In Model 1, we found genetic variants associated with increased EA, not controlling for CP, associated with decreased risk of suicide attempt (Model 1 SVMR OR, 0.524, 95% CI, 0.412-0.666, *P* = 1.07×10^−7^), and genetic variants associated with increased CP, not controlling for EA, also associated with decreased risk of suicide attempt (Model 1 SVMR OR, 0.714, 95% CI, 0.577-0.885, *P* = 0.002). In Model 2, accounting for comorbid mental disorders, we found associations of greater magnitude than in Model 1 but less precision for both EA (Model 2 SVMR OR, 0.687, 95% CI, 0.577-0.885, *P* = 0.002) and CP (Model 2 SVMR OR, 0.794, 95% CI, 0.636-0.990, *P* = 0.040) (Table 1, lines 1, 2).

### MVMR

We used MVMR to simultaneously assess the direct independent effects of each exposure, controlling for the other. In Model 1, controlling for CP, we found increased EA still associated with decreased risk of suicide attempt, with a greater magnitude for the point estimate but within the 95% confidence interval of the SVMR point estimate (Model 1 MVMR OR, 0.450, 95% CI, 0.314-0.644, *P* < 1E-04). Conversely, controlling for EA, we found CP no longer associated with suicide attempt risk (Model 1 MVMR OR, 1.044, 95% CI, 0.764-1.426, *P* = 0.786).

Further adjusting Model 1 for income, we found increased EA still associated with decreased risk of suicide attempt: the point estimate was greater, and no longer within the 95% confidence interval of the SVMR point estimate (Model 1 OR, 0.342, 95% CI, 0.206-0.568, *P* = 1.61⨯10^−4^); CP, adjusting for income as well as EA, is still no longer associated directly with suicide attempt risk (Model 1 OR, 1.182, 95% CI, 0.842-1.659, *P* = 0.333) (Table 1, lines 3,4; compared to lines 1, 2).

These results appear to be robust: In Model 2, accounting for comorbid mental disorders, as well as adjusting for income, we found significant associations of increased EA with decreased suicide attempt risk of similar magnitude and within the 95% confidence interval of Model 1 (Model 1 OR, 0.342, 95% CI, 0.206-0.568, *P* = 1.61×10^−4^; Model 2 OR, 0.450, 95% CI, 0.314-0.644, *P* < 1E-04); CP still not significant (Model 1 OR, 1.182, 95% CI, 0.842-1.659, *P* = 0.333; Model 2 OR, 1.143, 95% CI, 0.803-1.627, *P* = 0.475) (See Table 1, lines 6, 7).

### Sensitivity analysis

The MR PRESSO global tests and Cochran Q tests did not indicate heterogeneity, nor did the MR Egger intercepts indicate directional pleiotropy in any of the analyses, after outliers identified by MR PRESSO were removed as sources of potential pleiotropy.

## DISCUSSION

In the current study, we use MR methods to highlight an important possible public health benefit of increasing EA: reducing the risk of suicidal behavior. Using multivariable MR, we found EA reduces the risk for having attempted suicide by approximately 65%, which supports of a causal pathway between EA and suicide attempt that is independent of cognitive performance or socioeconomic status, as measured by income. Further, this relationship remained significant in our additional analysis using a suicide attempt GWAS that accounts for psychiatric disorders.

We also found an inverse relationship between CP and suicide attempts that appears to be mediated through EA since the effect of CP on suicide attempt in our SVMR analysis was no longer significant after incorporating EA in the MVMR. Consistency of the EA estimates across complementary MR methods used to assess sensitivity to of these estimates to changes in the underlying IV assumptions also strengthens our inference of causality.

This study contributes to the burgeoning number of MVMR studies reporting an effect of EA on health outcomes after accounting for cognitive performance [31, 34, 53, 54], which given the disproportionate disparity in suicide rates between individuals with a high school education or less compared to college graduates [15, 18] suggests a contribution to the substantial health inequalities observed associated with differences in EA [55]. Other study designs using innovative approaches have also found evidence for causal effects of education on health outcomes [55]. For example, natural experiments, like compulsory educational reforms, that take advantage of external changes that affect EA but are unrelated to health, have shown significant causal effects on general health outcomes [56, 57], while twin design studies have found similarly strong causal effects on health outcomes [58] and mortality [59].

Notably, in our single-variable MR analysis, we found that CP reduces the risk for attempting suicide by almost 30%; however, after incorporating EA and income into a MVMR framework, CP was no longer significantly associated with suicidal behavior, which supports previous observational literature finding that EA explains a substantial proportion of the association between CP and suicidal behavior [60-62]. CP and EA are genetically correlated [36], and there is MR evidence supporting a bi-directional relationship between EA and CP [54]; however, using multivariable MR methods and sensitivity analyses to assess for possible pleiotropy, we were able to extricate their direct effects on the risk for having attempted suicide, which is critical when considering policies aimed at preventing suicide. Taken together, our results lend support to the notion that programs aimed at increasing the years of schooling may reduce the increasing age-adjusted suicide rates. There may be other added benefits of increases EA, with schools as ideal settings for therapeutic interventions including dialectical and cognitive-behavioral therapy [63].

MR does not elucidate the pathways between environmental exposures and outcomes [53], but possible mediating mechanisms include economic, health-behavioral (smoking, alcohol consumption, diet, etc.), psychosocial (e.g., social support networks to deal with daily stressors [64]), and access to health care [65]. Smoking has been linked with increased suicide risk [66]; however, a recent univariable MR found no evidence that smoking directly affects suicide risk [67]. Problematic alcohol consumption may be one of the many possible health-behavioral pathways linking EA and suicide attempts. While the association between alcohol dependence and suicide has been known for decades [68], we recently reported an MR study that found evidence EA causally impacts drinking patterns and the risk for AD with increased EA reducing the frequency of binge drinking ≥6 drinks per occasion, and the risk for AD by about 50% [28]. Further, alcohol is often heavily consumed prior to suicide [69, 70] and Kaplan et al., (2014) showed, after controlling for problem drinking behavior, suicide completers had a 140% increased risk for alcohol consumption prior to death [71]. Differences in the prevalence of risk factors like alcohol consumption likely work in combination with, for example, limitations in access and differences in the ability to benefit from health care and medical information to contribute to the widening educational gradient in mortality [15, 72, 73].

## STRENGTHS & LIMITATIONS

There are notable strengths of the current study, including the use of multiple MR sensitivity analyses (Egger, weighted median, and weighted mode MR) that each accommodate different assumptions about genetic pleiotropy to test the robustness of the IVW estimate [74]. In addition, the GWAS summary statistics of suicide attempt employed as our outcome are based on hospital records, and not self-reported suicide attempt, which may be prone to social-desirability and recall biases [75]. Similarly, while 90% of those attempting suicide have at least one psychiatric disorder [8], our analyses included examining the relationship between EA and suicide attempt using models accounting DSM-IV psychiatric disorders [40], which improves the generalizability of the our findings.

Interpreting the results of this study requires an understanding of its limitations. While MVMR enables the simultaneous assessment of the effects of two or more exposures, other horizontal pleiotropy pathways, like personality traits, may still bias the estimates [31]. However, using different sensitivity analyses that have orthogonal assumptions to test for potential pleiotropy [46], we found similar estimates across the methods. Next, it has been shown that genetic variants associated with EA are also linked with family background and parental EA [76] suggesting residual population stratification, assortative mating, and dynastic effects – where parental EA and cognitive performance affect the life outcomes of their offspring – could potentially explain these findings. Also, our EA instrument only evaluated the number of years of schooling at academic institutions, so resolving which aspects of education, or even how skills and values learned outside of formal academic training, impact suicide risk will need to be evaluated by future studies. Similarly, cognitive performance represents and amalgamation of functions responsible for perception, thought, action, and emotion [34, 77], which might explain different aspects of the progression from suicidal ideation to attempts to completion [60].

Therefore, given the complexity cognitive performance [78] and the possible differential effects various components of cognitive performance have on suicide risk, the verbal-numerical attainment and neuropsychological tests used to generate the cognitive performance outcome in the Lee et al. (2018) GWAS is insufficient to elucidate these differences. Therefore, future studies disentangling the role of distinct aspects of cognitive performance in suicidal behavior are necessary.

Also, given the included datasets were of primarily European Ancestry, participants in the UK Biobank more educated, leads healthier lifestyles, and have fewer health problems than the general UK population [79], and that it was recently shown that EA protected non-Hispanic Whites, but not non-Hispanic Blacks against future suicide attempts and deaths [80], future studies replicating these results in other ethnic populations are warranted. Further, the nature of suicidality makes studying suicidal behavior difficult [81], and while suicide attempts are strong predictors of later suicide completions with 25% of those dying by suicide having a failed attempt within the year prior to completion [3], we were unable to differentiate between violent and non-violent suicide attempts which confer significantly different risks for later completions [82, 83]. Further still, we were unable to distinguish the effects of gender: sex-specific suicide attempt GWASs are not yet available. However, gender and age were covariates in the GWAS logistic regression, so the outcome phenotype would be interpreted as risk of suicide attempt controlling for gender. Finally, we were unable to assess suicidal behavior within the ideation- to-action framework where suicidal ideation, the progression from ideation to attempts, are postulated to be distinct phenomena with unique predictors [81], which makes future MR studies using GWAS that differentiate between ideation and attempts necessary when the data becomes available.

## CONCLUSION

This study was, to our knowledge, the first MR study to examine the effects of EA and CP on suicidal behaviors. Supporting observational studies, and under MR assumptions, our MVMR analyses suggest that EA causally impacts the risk for suicide attempt, while the negative effect of CP on risk of suicide attempt may be mediated through the effect of CP on EA, which together suggests that targeted prevention programs that increase EA may be useful to reduce the mortality due to suicide.

## Data Availability

All analyses were conducted using publicly available data. The summary-level data for educational attainment and cognitive performance are available at https://www.thessgac.org/data; the summary level data for income (MRC-IEU UK Biobank GWAS Pipeline) are available through MR Base at http://www.mrbase.org/; the summary level data for suicide attempt (iPSYCH) are available at https://ipsych.dk/forskning/downloads/

## ACKNOWLEGEMENTS

This research was facilitated by the Social Science Genetic Association Consortium (SSGAC), The Lundbeck Foundation Initiative for Integrative Psychiatric Research (iPSYCH), and the Medical Research Council Integrative Epidemiology Unit (MRC-IEU, University of Bristol, UK), especially the developers of the MRC-IEU UK Biobank GWAS Pipeline. We gratefully acknowledge their contributing studies and the participants in those studies without whom this effort would not be possible. This work was supported by the National Institutes of Health (NIH) intramural funding [ZIA-AA000242 to F.W.L]; Division of Intramural Clinical and Biological Research of the National Institute on Alcohol Abuse and Alcoholism (NIAAA). The authors declare no conflict of interest.

## CONFLICT OF INTEREST

The authors declare no conflict of interest.

## CODE AVAILABILITY

The analysis code in R is available upon request and all data displayed in the figures are available in the Supplementary tables.

## Notes

### Competing Interest Statement

The authors have declared no competing interest.

## REFERENCES

1. Collaborators, G.M.a.C.o.D., Global, regional, and national life expectancy, all-cause mortality, and cause-specific mortality for 249 causes of death, 1980-2015: a systematic analysis for the Global Burden of Disease Study 2015. Lancet, 2016. 388(10053): p. 1459–1544.

2. Hedegaard, H., S.C. Curtin, and M. Warner, Suicide Mortality in the United States, 1999-2017. NCHS Data Brief, 2018(330): p. 1–8.

3. Owens, D., J. Horrocks, and A. House, Fatal and non-fatal repetition of self-harm: Systematic review. British Journal of Psychiatry, 2002. 181(3): p. 193–199.

4. Curtis Florence, T.S., Tamara Haegerich, Feijun Luo, Chao Zhou. Estimated Lifetime Medical and Work-Loss Costs of Fatal Injuries — United States, 2013. Morbidity and Mortality Weekly Report (MMWR) 2015 October 2, 2015 [cited 2019 November 9]; Available from: https://www.cdc.gov/mmwr/preview/mmwrhtml/mm6438a4.htm.

5. Ludwig, B., et al., The Life Span Model of Suicide and Its Neurobiological Foundation. Frontiers in neuroscience, 2017. 11: p. 74–74.

6. Ports, K.A., et al., Adverse Childhood Experiences and Suicide Risk: Toward Comprehensive Prevention. American Journal of Preventive Medicine, 2017. 53(3): p. 400–403.

7. Olfson, M., et al., National Trends in Suicide Attempts Among Adults in the United States. JAMA Psychiatry, 2017. 74(11): p. 1095–1103.

8. Mullins, N., et al., GWAS of Suicide Attempt in Psychiatric Disorders and Association With Major Depression Polygenic Risk Scores. Am J Psychiatry, 2019. 176(8): p. 651–660.

9. Mirkovic, B., et al., Genetic Association Studies of Suicidal Behavior: A Review of the Past 10_jYears, Progress, Limitations, and Future Directions. Frontiers in psychiatry, 2016. 7: p. 158–158.

10. Kendler, K.S., Genetic and environmental pathways to suicidal behavior: reflections of a genetic epidemiologist. Eur Psychiatry, 2010. 25(5): p. 300–3.

11. Brent, D.A. and N. Melhem, Familial transmission of suicidal behavior. Psychiatr Clin North Am, 2008. 31(2): p. 157–77.

12. Strawbridge, R.J., et al., Identification of novel genome-wide associations for suicidality in UK Biobank, genetic correlation with psychiatric disorders and polygenic association with completed suicide. EBioMedicine, 2019. 41: p. 517–525.

13. Crosby, A.E., L. Ortega, and M.R. Stevens, Suicides - United States, 2005-2009. MMWR Suppl, 2013. 62(3): p. 179–83.

14. Lorant, V., et al., Socio-economic inequalities in suicide: a European comparative study. Br J Psychiatry, 2005. 187: p. 49–54.

15. Phillips, J.A. and K. Hempstead, Differences in U.S. Suicide Rates by Educational Attainment, 2000-2014. Am J Prev Med, 2017. 53(4): p. e123–e130.

16. Abdel-Rahman, O., Socioeconomic predictors of suicide risk among cancer patients in the United States: A population-based study. Cancer Epidemiol, 2019. 63: p. 101601.

17. Olshansky, S.J., et al., Differences in life expectancy due to race and educational differences are widening, and many may not catch up. Health Aff (Millwood), 2012. 31(8): p. 1803–13.

18. Case, A. and A. Deaton, Rising morbidity and mortality in midlife among white non-Hispanic Americans in the 21st century. Proceedings of the National Academy of Sciences, 2015. 112(49): p. 15078.

19. Leamer, E.E., Let’s Take the Con Out of Econometrics. The American Economic Review, 1983. 73(1): p. 31–43.

20. Smith, G.D. and S. Ebrahim, ‘Mendelian randomization’: can genetic epidemiology contribute to understanding environmental determinants of disease? Int J Epidemiol, 2003. 32(1): p. 1–22.

21. Smith, G.D. and S. Ebrahim, Epidemiology - is it time to call it a day? International Journal of Epidemiology, 2001. 30(1): p. 1–11.

22. Phillips, A.N. and G.D. Smith, How Independent Are Independent Effects - Relative Risk-Estimation When Correlated Exposures Are Measured Imprecisely. Journal of Clinical Epidemiology, 1991. 44(11): p. 1223–1231.

23. Westman, J., et al., The influences of place of birth and socioeconomic factors on attempted suicide in a defined population of 4.5 million people. Arch Gen Psychiatry, 2003. 60(4): p. 409–14.

24. Qin, P., E. Agerbo, and P.B. Mortensen, Suicide risk in relation to socioeconomic, demographic, psychiatric, and familial factors: a national register-based study of all suicides in Denmark, 1981-1997. Am J Psychiatry, 2003. 160(4): p. 765–72.

25. Bothwell, L.E., et al., Assessing the Gold Standard — Lessons from the History of RCTs. New England Journal of Medicine, 2016. 374(22): p. 2175–2181.

26. Gunnell, D., A Population Health Perspective on Suicide Research and Prevention. Crisis, 2015. 36(3): p. 155–60.

27. Smith, G.D., Use of genetic markers and gene-diet interactions for interrogating population-level causal influences of diet on health. Genes and Nutrition, 2011. 6(1): p. 27–43.

28. Rosoff, D.B., et al., Educational attainment impacts drinking behaviors and risk for alcohol dependence: results from a two-sample Mendelian randomization study with ∼780,000 participants. Molecular Psychiatry, 2019.

29. Gage, S.H., et al., Investigating causality in associations between education and smoking: a two-sample Mendelian randomization study. International Journal of Epidemiology, 2018. 47(4): p. 1131–1140.

30. Sanderson, E., et al., An examination of multivariable Mendelian randomization in the single-sample and two-sample summary data settings. Int J Epidemiol, 2019. 48(3): p. 713–727.

31. Davies, N.M., et al., Multivariable two-sample Mendelian randomization estimates of the effects of intelligence and education on health. Elife, 2019. 8.

32. Carter, A.R., et al., Understanding the consequences of education inequality on cardiovascular disease: mendelian randomisation study. BMJ, 2019. 365: p. 1855.

33. Tillmann, T., et al., Education and coronary heart disease: mendelian randomisation study. Bmj-British Medical Journal, 2017. 358.

34. Gill, D., et al., Education protects against coronary heart disease and stroke independently of cognitive function: evidence from Mendelian randomization. International Journal of Epidemiology, 2019.

35. Okbay, A., et al., Genome-wide association study identifies 74 loci associated with educational attainment. Nature, 2016. 533(7604): p. 539–42.

36. Lee, J.J., et al., Gene discovery and polygenic prediction from a genome-wide association study of educational attainment in 1.1 million individuals. Nature Genetics, 2018. 50(8): p. 1112–1121.

37. Deary, I.J., et al., Intelligence and educational achievement. Intelligence, 2007. 35(1): p. 13–21.

38. Deary, I.J., L. Penke, and W. Johnson, The neuroscience of human intelligence differences. Nature Reviews Neuroscience, 2010. 11(3): p. 201–211.

39. Burgess, S. and S.G. Thompson, Multivariable Mendelian randomization: the use of pleiotropic genetic variants to estimate causal effects. Am J Epidemiol, 2015. 181(4): p. 251–60.

40. Erlangsen, A., et al., Genetics of suicide attempts in individuals with and without mental disorders: a population-based genome-wide association study. Mol Psychiatry, 2018.

41. Morris, T.T., N.M. Davies, and G. Davey Smith, Can education be personalised using pupils’ genetic data? bioRxiv, 2019: p. 645218.

42. Hill, W.D., et al., A combined analysis of genetically correlated traits identifies 187 loci and a role for neurogenesis and myelination in intelligence. Molecular Psychiatry, 2019. 24(2): p. 169–181.

43. Elsworth, B., Mitchell, R, Raistrick, CA, Paternoster, L, Hemani, G, Gaunt, TR MRC IEU UK Biobank GWAS pipeline version 1. 2017. DOI: https://doi.org/10.5523/bris.2fahpksont1zi26xosyamqo8rr.

44. Palmer, T.M., et al., Using multiple genetic variants as instrumental variables for modifiable risk factors. Stat Methods Med Res, 2012. 21(3): p. 223–42.

45. Burgess, S., N.M. Davies, and S.G. Thompson, Bias due to participant overlap in two-sample Mendelian randomization. Genetic epidemiology, 2016. 40(7): p. 597–608.

46. Hemani, G., et al., The MR-Base platform supports systematic causal inference across the human phenome. Elife, 2018. 7.

47. Bowden, J., G.D. Smith, and S. Burgess, Mendelian randomization with invalid instruments: effect estimation and bias detection through Egger regression. International Journal of Epidemiology, 2015. 44(2): p. 512–525.

48. Davey Smith, G., et al., Assessing the suitability of summary data for two-sample Mendelian randomization analyses using MR-Egger regression: the role of the I2 statistic. International Journal of Epidemiology, 2016. 45(6): p. 1961–1974.

49. Rees, J.M.B., A.M. Wood, and S. Burgess, Extending the MR-Egger method for multivariable Mendelian randomization to correct for both measured and unmeasured pleiotropy. Stat Med, 2017. 36(29): p. 4705–4718.

50. Bowden, J., et al., A framework for the investigation of pleiotropy in two-sample summary data Mendelian randomization. Statistics in Medicine, 2017. 36(11): p. 1783–1802.

51. Bowden, J., et al., Improving the accuracy of two-sample summary-data Mendelian randomization: moving beyond the NOME assumption. Int J Epidemiol, 2019. 48(3): p. 728–742.

52. Verbanck, M., et al., Detection of widespread horizontal pleiotropy in causal relationships inferred from Mendelian randomization between complex traits and diseases. Nat Genet, 2018. 50(5): p. 693–698.

53. Sanderson, E., et al., Mendelian randomisation analysis of the effect of educational attainment and cognitive ability on smoking behaviour. Nature Communications, 2019. 10(1): p. 2949.

54. Anderson, E.L., et al., Education, intelligence and Alzheimer’s disease: Evidence from a multivariable two-sample Mendelian randomization study. bioRxiv, 2018: p. 401042.

55. Zajacova, A. and E.M. Lawrence, The Relationship Between Education and Health: Reducing Disparities Through a Contextual Approach. Annu Rev Public Health, 2018. 39: p. 273–289.

56. Fletcher, J.M., New evidence of the effects of education on health in the US: compulsory schooling laws revisited. Social science & medicine (1982), 2015. 127: p. 101–107.

57. Davies, N.M., et al., The causal effects of education on health outcomes in the UK Biobank. Nature Human Behaviour, 2018. 2(2): p. 117–125.

58. Gerdtham, U.G., et al., Do Education and Income Really Explain Inequalities in Health? Applying a Twin Design. The Scandinavian Journal of Economics, 2016. 118(1): p. 25–48.

59. Lundborg, P., C.H. Lyttkens, and P. Nystedt, The Effect of Schooling on Mortality: New Evidence From 50,000 Swedish Twins. Demography, 2016. 53(4): p. 1135–1168.

60. Hansson Bittár, N., D. Falkstedt, and A. Sörberg Wallin, How intelligence and emotional control are related to suicidal behavior across the life course – A register-based study with 38-year follow-up. Psychological Medicine: p. 1–7.

61. Sorberg, A., et al., Cognitive ability in early adulthood is associated with later suicide and suicide attempt: the role of risk factors over the life course. Psychol Med, 2013. 43(1): p. 49–60.

62. Sorberg Wallin, A., et al., Childhood IQ and mortality during 53 years’ follow-up of Swedish men and women. J Epidemiol Community Health, 2018. 72(10): p. 926–932.

63. Evans, R. and C. Hurrell, The role of schools in children and young people’s self-harm and suicide: systematic review and meta-ethnography of qualitative research. BMC Public Health, 2016. 16(1): p. 401.

64. Taylor, S.E. and T.E. Seeman, Psychosocial resources and the SES-health relationship. Ann N Y Acad Sci, 1999. 896: p. 210–25.

65. Cutler, D. and A. Lleras-Muney, Education and Health: Evaluating Theories and Evidence, in Making Americans Healthier: Social and Economic Policy as HealthPolicy,

66. J. House, et al., Editors. 2008, Russell Sage Foundation: New York. Poorolajal, J. and N. Darvishi, Smoking and Suicide: A Meta-Analysis. PloS one, 2016. 11(7): p. e0156348–e0156348.

67. Harrison, R., et al., Examining the effect of smoking on suicidal ideation and attempts: A triangulation of epidemiological approaches. medRxiv, 2019: p. 19007013.

68. Pompili, M., et al., Suicidal behavior and alcohol abuse. International journal of environmental research and public health, 2010. 7(4): p. 1392–1431.

69. Conner, K.R., et al., Acute use of alcohol and methods of suicide in a US national sample. American journal of public health, 2014. 104(1): p. 171–178.

70. Kaplan, M.S., et al., Acute alcohol intoxication and suicide: a gender-stratified analysis of the National Violent Death Reporting System. Inj Prev, 2013. 19(1): p. 38–43.

71. Kaplan, M.S., et al., Use of alcohol before suicide in the United States. Annals of epidemiology, 2014. 24(8): p. 588-592.e5922.

72. Lleras-Muney, A., The Relationship between Education and Adult Mortality in the United States. The Review of Economic Studies, 2005. 72(1): p. 189–221.

73. Meara, E.R., S. Richards, and D.M. Cutler, The gap gets bigger: changes in mortality and life expectancy, by education, 1981-2000. Health affairs (Project Hope), 2008. 27(2): p. 350–360.

74. Lawlor, D.A., Commentary: Two-sample Mendelian randomization: opportunities and challenges. International Journal of Epidemiology, 2016. 45(3): p. 908–915.

75. Choi, B.C. and A.W. Pak, A catalog of biases in questionnaires. Prev Chronic Dis, 2005. 2(1): p. A13.

76. Kong, A., et al., The nature of nurture: Effects of parental genotypes. Science, 2018. 359(6374): p. 424–428.

77. Deary, I.J. and W. Johnson, Intelligence and education: causal perceptions drive analytic processes and therefore conclusions. Int J Epidemiol, 2010. 39(5): p. 1362–9.

78. Trampush, J.W., et al., GWAS meta-analysis reveals novel loci and genetic correlates for general cognitive function: a report from the COGENT consortium. Molecular Psychiatry, 2017. 22(3): p. 336–345.

79. Fry, A., et al., Comparison of Sociodemographic and Health-Related Characteristics of UK Biobank Participants With Those of the General Population. American Journal of Epidemiology, 2017. 186(9): p. 1026–1034.

80. Assari, S., et al., Higher Educational Attainment is Associated with Lower Risk of a Future Suicide Attempt Among Non-Hispanic Whites but not Non-Hispanic Blacks. J Racial Ethn Health Disparities, 2019. 6(5): p. 1001–1010.

81. Klonsky, E.D., A.M. May, and B.Y. Saffer, Suicide, Suicide Attempts, and Suicidal Ideation. Annu Rev Clin Psychol, 2016. 12: p. 307–30.

82. Runeson, B., et al., Method of attempted suicide as predictor of subsequent successful suicide: national long term cohort study. BMJ, 2010. 341: p. c3222.

83. Stefansson, J., P. Nordstrom, and J. Jokinen, Suicide Intent Scale in the prediction of suicide. J Affect Disord, 2012. 136(1-2): p. 167–171.

